# Care home’s resilience during the pandemic: changing to adapt

**DOI:** 10.1101/2023.11.23.23298949

**Authors:** Miguel Vasconcelos Da Silva, Zunera Khan, Lise Birgitte Austbø Holteng, Martha Therese Gjestsen, Dag Aarsland, Ingelin Testad

## Abstract

**Introduction:** The unfolding of the pandemic in 2020 led to an unprecedented level of stress in care homes, including adjustments and changes within this pivotal service to the community in coping with the increased pressures on and need of support for the National Health Service (NHS). Care homes (CH) were crucial in providing healthcare services, but the support given to them was limited and lacked strategic information that enabled them to experience better outcomes. It was important to understand the CH’s views and strategies and how these endured during the pandemic.

**Methods:** A total of 220 CH from the Care Home Research Network (CHRN) were invited to take part in an on-line survey, where 51 CH completed the survey. The survey comprised demographic- and open-ended questions concerning the service and adaptations/adjustments made within CH during the pandemic.

**Results:** CH staff reported an increase in the level of stress in their roles, including their workload, due to COVID-19, which made it difficult to cope with their tasks. CH also reported the fear of uncertainty, with some CH saying there was a lack of information. Loneliness and depression were reported as colossally increasing for residents. CH made changes to their working ways and spaces, adopting a more frequent use of technology platforms to meet residents’ needs.

**Conclusion:** CH were resilient and adaptable to highly stressful challenges, ensuring residents’ need were met. CH felt a huge pressure to support wider health services, whilst having to deal with uncertainty, staff and supply shortages as well as cope with the loss of residents. Nevertheless, CH reinvented themselves, promoted more teamwork and used supporting channels, including investment in technology to facilitate communication between residents’ and families but also with supporting services such as general practitioners (GPs). Despite these changes, loneliness and depression amongst residents was reported as high. Highlighting the need to assess the long-term impacts of this on residents includes the development of interventions or strategies that will reduce depression/loneliness.

## INTRODUCTION

The spread of COVID-19 in the UK caused considerable challenges across all areas of society; the health and social care system particularly faced a huge additional strain, impacting care staff, residents and their families [1]. During the onset of the pandemic between 2020 and 2021, the government imposed many restrictions to slow the spread of the virus and minimise the impact on the National Health Service (NHS) [2].

People over the age of 70 and people with certain health conditions (i.e., diabetes, respiratory diseases) were at increased risk of developing severe illness and death from COVID-19; therefore, extra measures, like self-isolation and social distancing, were set by the government to protect them [3].

Approximately 420,000 of the UK elderly population live within the 15□487 care homes (CH): residential homes (11□333) and nursing homes (4413) registered with the Care Quality Commission (CQC) [4]. The majority of people living in CH are elderly, aged above 70, having multiple comorbidities and thus at higher risk of severe illness and death from COVID-19, along with the possibility of increasing feelings of loneliness, agitation and low mood due to the extra measures [3].

CH are funded through a complex mix of sources (self-funding, local authorities and healthcare funding from NHS), and they are not part of the NHS. They are run by independent organisations, with large for-profit organisations comprising a third of the providers and the remainder consisting of not-for-profit third-sector organisations, or small private companies with only a small number of homes.

Historically, CH lacked the necessary funding and support to run smoothly [5], and with the surge of the pandemic, CH needed more support to be able to meet the increased demands and care needs and cope with the pandemic challenges. CH staff worked relentlessly to cope with the pandemic and had to be more resourceful in order to meet the needs of residents, which was challenging and frequently brought them into the news spotlight during the pandemic. In addition, CH received extra pressure in supporting health services by admitting patients from hospitals [6]. Furthermore, the number of excess deaths CH experienced also meant additional work for care staff who had to provide more end-of-life care even though training was limited.

The government released specific guidance on residential care provision, including stopping all visits to residents from family and friends [7]. Feelings of loneliness and isolation are common among CH residents and the elderly [8], and this is likely to increase with restrictions on visitors and social activities [9]. There is growing evidence to suggest that social isolation and loneliness are risk factors for both physical and mental illness [10-12]. Loneliness is also associated with an increased risk of dementia [13]. It is therefore important to understand the restrictions imposed during the COVID-19 pandemic, such as social distancing measures, and the impact these had on the well-being of residents and staff. It is also important to examine what strategies CH used to minimise this, and whether these were effective.

Whilst there were strict social distancing measures in place, CH needed to implement other ways for residents to feel connected, have meaningful interactions, and ensure the optimal management of health conditions, symptoms and concerns. Previous research on the use of video calls and telephone befriending have demonstrated that these are ways to increase meaningful interactions between a resident and family member who is no longer allowed to visit them. However, it is unknown whether CH routinely utilise these tools, and whether there were sufficient resources to implement them during the crisis [14, 15].

Moreover, CH are reliant on GP services to meet residents’ medical needs, and residential homes with no onsite nursing expertise also rely on community health care services to meet residents’ health needs. There is limited understanding of how integrated work between health and social care during the COVID-19 pandemic responded to meet the increasing needs of older people in care home and ensure care home staff were supported. It is therefore imperative to collect data on the strategies needed to maintain social interactions among residents, facilitate integration between health and social care staff for optimal management of symptoms and concerns, and assess whether these measures have been effective. If so, how resource intensive these are, and whether it is feasible to roll them out on a larger scale in other care home, should be explored.

It is important to understand how CH managed the additional burden to staff and residents and assess whether these methods were effective in order to provide evidence-based guidelines during this pandemic for future health crises. Additionally, there is a need to understand the impact of COVID-19 in the care home setting and how strategies have been effective in preventing spread of the virus. It is also likely that, even though restrictions have been lifted, we will continue to see the effects on the psychological and physical health of care home residents and care staff. Therefore, it is essential to develop effective measures to reduce this impact and minimise poor health outcomes related to COVID-19.

This study seeks to explore how the pandemic impacted CH and how they coped and adapted to working in the context of the pandemic by providing a first-hand opinion from the CH. The study also helps to share CH’s perspectives, experiences, and strategies during the pandemic.

## METHODS

This was a mixed methods study, which included quantitative data and qualitative data with an inductive approach, aiming to explore the experience of CH during the pandemic. An online open survey was launched, and 220 CH, members of the Care Home Research Network King’s College London and Maudsley BRC, were invited to take part in the study; responses were anonymous. The survey comprised questions about CH demographics and explored their views on the impact that the pandemic had on CH staff and residents, including questions about the challenges they faced and how they managed or adapted to working during the pandemic. Ethical Clearance was obtained from King’s College London Research Ethics Office (Ethical Clearance Reference Number: LRS-19/20-19657). Participants’ consent was obtained online before their completion of the anonymous survey.

Data analysis was completed using systematic text condensation as described by Kirsti Malterud [16], based on Giorgio’s psychological phenomenological analysis. The aim of the phenomenological analysis is to develop knowledge about the participants’ experiences [16]. Phenomenology focuses on studying a phenomenon from the perspective of the individual. It is important to approach the phenomenon with an open mind to ensure that the topic of research is not sculpted into predetermined categories, hence this approach emphasises a contextualised description that is close to what was experienced [16].

Reports were read through by the researchers to obtain an overall impression. After reading the reports several times, the researchers agreed on some preliminary main themes. Meaning units were then identified and classified into codes, and the decontextualised material was discussed by the researchers. The codes and meaning units were rearranged several times before subgroups were created for each code group to view important aspects. Condensation and abstraction of the meaning units within each code and writing artificial quotes was then performed. Finally, content from the previous steps was re-contextualised into an analytical text with a category heading. Artificial quotes from the third step were rewritten into analytical text according to the results’ main themes and assembled with a quote that was representative of the theme.

## Results

### Care Home demographics

Fifty-one CH responded to the survey, with the majority of respondents occupying the role of manager. The type of home was divided into Nursing Home and Residential and Care Home, with most CH (61%, n= 31) having between 30 to 100 beds and 69% (n=35) a CQC rating of good, see Table 1.

**Table 1.** respondents and CH demographics

| Number Of CH responses | Responding role | % | Type of unit | % | Number of beds | % | Staff | % | CQC rating | % |
| --- | --- | --- | --- | --- | --- | --- | --- | --- | --- | --- |
| 51 | Manager | 94% | Nursing Home | 33% | <30 | 31% | <30 | 31% | Requires improvement | 27% |
|  | Clinical lead | 2% | Residential | 31% | 30-100 | 61% | 30-100 | 55% | Good | 69% |
|  | Deputy | 2% | Care home | 37% | >100 | 8% | >100 | 12% | outstanding | 4% |
|  | Coordinator | 2% |  |  |  |  |  |  |  |  |

**Table 2.** frequency of the COVID-19 overall impact.

| Care home affected by pandemic | % | Residents affected | % | Residents passed away due to COVID-19 | % | Admissions to care homes with COVID-19 | % | Staff sickness due to COVID-19 | % |
| --- | --- | --- | --- | --- | --- | --- | --- | --- | --- |
| Yes | 78% | yes | 61% | yes | 53% | yes | 20% | yes | 76% |
| no | 22% | no | 35% | no | 45% | no | 72% | no | 20% |
|  |  | No response | 4% | No response | 3% | Prefer not to answer | 8% | Prefer not to answer | 4% |

**Table 3.** Example from the analysis step 2, 3 and 4.

| **Preliminary theme** | **Meaning Unit** | **Artificial quote - condensate** | **Analytical text** |
| --- | --- | --- | --- |
| Emotional strain | *"The fact that we were not able to have visitors; the fear that was felt by staff; fear of the unknown."* | The participants said that they were afraid of the consequences of the pandemic, the isolation and the uncertainty. | Feelings of fear about the situation and the uncertainty, fear of infection, the negative impact of loneliness and losing their care home residents greatly affected the staff. They report that dealing with many deaths over a short period was very difficult, and watching their residents die in the aftermath of COVID-19 was highly strenuous. Some even reported that they were unsure about the future of the care home, and staff stopped coming to work because they were afraid of infection. |

### Impact on care homes

An increased level of stress in their roles due to COVID-19 was reported by 96% of the respondents, with 24% (n=12) saying that their workload had increased so much that they were unable to cope. Furthermore, staff absence had increased due to COVID-19 in 71% of CH, and 56% of respondents (n=27) said they had hired new staff to help cope with the increased workload. Overall, 94% (n= 48) reported that their work environment was more stressful and uncertain than before the pandemic started.

Worries about health and risks posed by COVID-19 affected staff according to 90% of interviewees, with 96% of CH able to test all of their staff for COVID-19. About 72% of CH had enough supply of PPE, 22% had enough until a new delivery, whilst 4% did not have enough supply and 2% preferred not to answer.

Information regarding COVID-19 was satisfactory for 88% of CH (n=44), whereas the rest of the care home felt that they had not received adequate information, and 20% said they had not received enough information to help manage the services during COVID-19. Twenty percent of the homes (n=10) said that their local Care Commissioning Group (CCG) was unable to provide all the support they needed, with a further 14% preferring not to answer and 67% saying that their local CCG was able to provide all the support they needed. Twenty percent of the respondents also said that there was a lack of specific guidelines to follow.

When asked about the impact of COVID-19 on residents, 54% of respondents said that, overall, their residents felt more anxious; 43% said that residents felt more distressed and agitated. Twenty-two percent said that, overall, their residents had experienced more psychotic symptoms, and 56% said that, overall, their residents felt more depressed with 71% saying their residents felt lonelier.

Six percent of respondents felt that there was an increase on the use of hypnotics, antipsychotics and antidepressants, whereas 37% of respondents said that they had to refer residents to a GP due to increased agitation, and 60% referred residents to a GP due to COVID-19 like symptoms.

Eighty-six percent of respondents said that, overall, families of residents were more anxious and/or worried, with 88% saying that they felt they could support and reassure families.

### Qualitative findings

The aim of the phenomenological analysis was to develop knowledge about the CH experiences during the pandemic. Four main themes were identified: (I) Emotional strain (II) Communication, and (III) Management and organisational strategies.

### Emotional strain

One of the most featured negative impacts of the pandemic is the emotional strain felt by staff in the situation. Fear, uncertainty, and feelings of abandonment were prominent.

#### Loneliness

Social distancing and isolation resulted in feelings of loneliness in both residents and staff. Most informants report a lack of visits and not seeing family members as the most negative impact of COVID-19 in CH. Staff increased supervision of the residents to counteract the negative impact of isolation. However, this resulted in an increased workload for staff. Staff increased 1:1 time with the residents and spent time with them in the afternoon, which usually would have been family time.

> *“Residents were unable to see their loved ones and feel the touch of their loved ones.” Fear*

Feelings of fear about the situation, uncertainty, fear of infection, the negative impact of loneliness and losing their care home residents greatly affected the staff. They report that dealing with many deaths over a short period was very difficult, and watching their residents die in the aftermath of COVID-19 was highly strenuous. Some even reported that they were unsure about the future of the care home, and staff stopped coming to work because they were afraid of infection.

> *“The fact that we were not able to have visitors; the fear that was felt by staff; fear of the unknown (…) I just do not know how I got through this period; I have had to work tirelessly with no break the whole period.”*

#### Appreciation and abandonment

CH felt increased support and recognition from families and were appreciated by stakeholders, for instance, receiving thank-you cards and kind words from the local community. Staff reported loyalty, commitment from colleagues, and understanding and support from residents and their families.

> *“There were encouragement and appreciation from relatives of the works which have been carried out by the staff in looking after their loved ones.”*

CH felt a lack of support and felt abandoned by the government. The lack of understanding of the challenges associated with keeping residents and staff safe and happy within the context of a pandemic with the horror stories in the media was problematic. CH also highlight the failure to recognise care home staff as essential workers as one of the most negative aspects of the pandemic and report that they felt forgotten. Misinformation in the public domain on the absence of essential resources, whilst at the same time functioning a protected cushion for NHS, made the situation difficult.

> *“Care homes were totally abandoned. All deliveries stopped. All outside agency support stopped, we felt totally abandoned.”*

### Communication

Finding new ways to communicate with family and colleagues was highlighted by most CH. Increased use of technology and strategic communication among staff and family interactions were areas of change during the pandemic.

#### Increased use of technology

Most CH describe extensive incorporation of technological solutions to communicate internally and with the outside world. CH described using technology as a way to deliver care and make social contact and often reported it as a primary communication channel within the care home setting.

> *“There was a better use of technology throughout the care home”*

Contact with GPs was done using technological solutions, where assessments and visits were done virtually. Staff reported that this worked well and said they experienced a better response from the general practitioner. Virtual services and improved online communication were mentioned as a positive impact of the pandemic, which led to predictability and sustainability.

> *“We have virtual reviews and assessments instead of people just turning up.” Communication with family and relatives*

Families were restricted from visiting and had to interact virtually with their relatives, and staff described new and creative ways to maintain resident-family commutation. Staff made sure they updated family members with the changes, including weekly newsletters, introducing a blog and purchasing iPads to allow residents video communication with their family. Staff reported increased communication with residents’ family members to update them about their loved ones.

> *“We have SKYPE, Facebook and the Magic Moment App that families can sign into to see and send messages to their loved ones.”*

#### Staff communication and teamwork

Staff well-being and support were highlighted, and various measures were taken to ensure adequate communication between colleagues. CH introduced blogs to reach out to staff and connected using group Google hangouts to keep in contact and ensure social interactions with each other. CH also recommended that others focus on support and information in similar situations.

> *“Share information and support each other in social media groups (…) and always communicate and discuss concerns, deal with issues on the same day.”*

When asked if the pandemic positively impacted CH, several informants said there was an increase in teamwork and that the staff were closer. Staff within CH worked together and with multidisciplinary teams, strengthening their resilience.

> *“We, all the staff, worked together and extended our hands in all areas to support ourselves and the residents.”*

### Management and organization strategies

Strategies to enable CH to work in the context of the pandemic were highly focused on infection control and social distancing to ensure safety and protect vulnerable residents and staff.

#### Infection control

Infection Control strategies to restrict infection spreading via physical control were widely used, e.g., furniture layout, single rooms, scheduling staff, increased cleaning, and testing of staff/visitors. CH made extensive changes to their premises in order to facilitate quarantine. Installing basins, having infection control audits, and cohorting were mentioned.

> *“New wash hand basin installed at the home entrance for everyone to wash their hands before entering the home. Staff uniforms were laundered on-site.”*

#### Social distancing and isolation

CH took measures to ensure social distancing between residents, often isolating the residents in their rooms. Staff tried to maintain a two-meter distance in the care setting, where possible, and implemented alternative ways of visits. Some CH report an increase in using the care home surroundings and gardens and allowing “window visits” from family.

> *“Residents were encouraged to stay in their room where possible.”*

## Discussion

The COVID-19 Pandemic brought unprecedented pressures and challenges to CH, with a high proportion of residents dying due to COVID-19, and care home staff having to deal with emotional coping alongside the increased workload, whilst at the same time admitting residents from hospital to support those services, despite staff shortages and increased stress on care home staff. Nonetheless, managers and CH staff showed their resilience in handling the challenges presented to them, whilst creating new ideas and ways of adaptive working to ensure the residents needs were met, with the main changes being in communication, emotional strain and management and organisational strategies.

When it comes to information regarding COVID-19, almost 1 in 3 CH felt that they had not received adequate or sufficient information to help them manage services. According to Rajan et al., government policies did not protect the CH as intended, and staff felt unsupported during the first wave of the pandemic [17]. This highlights the need to have a more centralised information system to keep CH and staff up to date with changes and guidelines to facilitate the delivery of a more efficient response, which was deemed critical for the dissemination of skills and knowledge in a study by Gray *et. al* [18].

According to CH, residents felt more anxious, more distressed, and agitated, which in turn led to CH having to refer residents to GP services and increased workload for care home staff. This could in part have been dealt with by determining whether there could have been better information sharing with care home. However, the frequently changing guidelines were difficult to follow even when they were in place [17]. Residents’ outcomes worsened with the pandemic [19], and given the increased workload and staff shortages, it reduces their ability to provide the usual standard of care [20], which in turn could have added to the increased levels agitation, anxiety, depression and loneliness. Discrepancies and irregularities in guidelines and care have led to a considerable negative outcomes for care home residents.

The pandemic brought emotional strain that impacted the residents, family members and staff. The current findings indicate that increased loneliness and depression experienced by care home residents, combined with increased workload and staff shortages, and compounded by the isolation measures (social distancing, isolation in single rooms, no visitors), possibly led to the overall worst health outcomes for the care home residents. Loneliness is a significant problem for people living in CH [21], and previous literature indicates that loneliness has a damaging impact on mental and physical health, and this could have been intensified by COVID-19 measures in this vulnerable group [22], thus highlighting the urgent need to develop and implement evidence-based interventions to create resilience for this group, which is already under represented in research [23].

Isolation and being unable to see their loved ones were one of the key challenges raised by respondents, which meant residents were restricted in experiencing personal contact and touch from their loved ones, losing the emotional support provided by their families, which may have a detrimental impact on their physical health (i.e., cardiovascular) and mental health [24]. The limited social contact may explain the high levels of loneliness and depression reported amongst residents, as social interactions can greatly affect quality of life, especially in care home residents living with dementia [25]. However, the data were only collected during the pandemic and did not focus on isolation and its impact on care home residents. Research on loneliness in care home residents and its impacts remain limited [26]. Hence, future studies should examine this longitudinally, examining its impact on residents and staff, in order to seek urgent interventions to address this.

Fear and uncertainty were described by care staff, possibly leading to increasing levels of stress, sickness, and a higher work absence amongst care home staff, which was also noted in another study [27]. A substantial number of care home residents are living with dementia, and working with this patient group is already associated with an increased risk of stress and burnout [28]. Moreover, the increased number of deaths meant staff had to cope with the loss of residents in such challenging times, whilst also suggesting a lack of support for care home staff in end-of-life care and raising important concerns for the mental health and wellbeing of care staff. In addition to this, for some care home managers, there was an increased level of anxiety and concern about the future of the care home as an organisation. The emotional strain was also felt by the families due to the restrictions on visiting and limiting the social and physical contact with their loved ones. However, several CH reported that relatives interacted virtually with their loved ones, still raising the concern for the homes that had limited access to these resources, in addition to families and residents who did not know how to use these technologies. Giebel *et al*. states that the pandemic had a dramatic impact on all people surrounding a care home, and that the inability to visit residents led to stress and grief in family members who felt guilty for placing their loved ones in CH [27].

Communication was a key theme reported by care home staff, which changed considerably during the pandemic given the need to adopt new ways of working, including the acquisition of Information Technology (IT) equipment.

Communication had an overall positive effect on the care home sector with the incorporation of more technology. This change enabled care home residents to communicate with families more easily and kept them connected and updated.

Furthermore, these changes helped communication between CH and GPs, facilitating the referrals and providing medical care for the residents’ needs through virtual reviews and assessments. In addition to this, there was better teamwork and engagement between internal multidisciplinary teams, which might have helped improve cohesiveness at an organisational level. In summary CH developed and adapted to form more sustainable ideas such as use of IT to build better communication with GP and families.

However, some CH might have lacked the financial resources and struggled to acquire new or more IT equipment to evaluate these new communications channels. This study did not assess the financial strength of the CH and/or the actual level of IT acquisition. Nevertheless, a previous report shows that community-based technology uptake is low [29, 30], meaning CH usually lack the IT resources to facilitate computer and systems access, accompanied by the fact that staff are generally deficient in digital skills [29]. Future studies could explore this more in addition to the financial differences between CH and the impacts of these factors on the delivery of care.

Management and organisation strategies was another theme that focused on the ways CH worked in the context of the pandemic; these included infection control and strict pandemic measures. In this theme, we observed how CH had to reinvent themselves, making changes and adjustments to the layout in order to comply with COVID-19 national guidelines whilst trying to meet the residents’ needs. For instance, most CH had to repurpose the space by using it in innovative ways (using gardens for visits) or changing spaces (making cleaning stations and staff launderettes).

### Impact

Loneliness remained a significant problem for people living in CH, and this was exacerbated by the isolation measures implemented during the pandemic. A key learning opportunity is the capacity to adapt and change quickly during the context of a pandemic with the adoption of existing systems or interventions that can help address the presented issues, more specifically the use of technology to improve communication and reduce isolation in residents, further highlighting the need for the development and implementation of evidence-based interventions to create more resilience amongst CH staff and residents.

## Conclusion

In summary, CH not only needed to address the social needs of the residents/families, they also had to consider the care home staff needs. Care home staff reported that they had to support each other more, sharing information and using social media groups to interact. They also reported the introduction of more staff wellbeing support and attending to their mental health needs.

It was a challenging time with increased loneliness and social isolation where residents, families and care home staff were highly impacted, yet it still brought some positive changes to CH: teams worked better and more cohesively, and families came closer with care home residents and staff, with better technology and communication. Overall, the current study reflects on key themes and ways through which CH proved their resilience and adaptiveness to challenges in order to meet residents’ needs and provision of care. The analysis and report on the ideas and solutions being implemented during pandemic can help guide best practices and models of care in addition to improving readiness and resilience for future national and international emergencies.

## Data Availability

All data produced in the present study are available upon reasonable request to the authors.

